# The Non-Linear Metabolic Architecture of Depression Risk: A Cross-Phenotypic Hybrid Modelling Analysis in Genes and Health Cohort

**DOI:** 10.64898/2026.11.24.26368843

**Authors:** Gaurav Thareja, Nicholas Bass, Gerome Breen

**Affiliations:** Social, Genetic, and Developmental Psychiatry Centre, Institute of Psychiatry, Psychology and Neuroscience, King’s College London, London, UK; Diagnostic Memory Clinic, East London NHS Foundation Trust, London, England, UK; Division of Psychiatry, University College London, London, England, UK

## Abstract

**Background:** Cardiometabolic dysregulation and depression frequently co-occur, yet studies often assume linear biomarker–depression relationships and under-represent South Asian populations. We investigated linear and non-linear associations between cardiometabolic biomarkers and electronic health record (EHR)-derived depression in British Bangladeshi and British Pakistani adults.

**Methods:** We analysed Genes C Health data across five EHR biomarkers: body mass index (BMI), glycated haemoglobin (HbA1c), high-density lipoprotein (HDL) cholesterol, low-density lipoprotein (LDL) cholesterol and triglycerides. In a primary development cohort (N = 6,340; 1,585 cases matched 1:3 to 4,755 controls), we applied principal component analysis and sex- and ethnicity-adjusted logistic regression to compare linear, restricted cubic spline, and optimised hybrid models. Models were validated externally in two temporal holdout sets sharing 18,733 controls: Holdout Set 1 (281 cases measured 6–12 months pre-diagnosis; N = 19,014) and Holdout Set 2 (1,815 cases measured >12 months pre-diagnosis; N = 20,548).

**Results:** Depression cases exhibited higher median HbA1c and lower LDL cholesterol in both ethnic groups, alongside higher BMI and triglycerides among Pakistani participants (all p < 0.05). The first three principal components explained 72.7% of metabolic variance with extensive ethnic overlap. Splines revealed non-linear trajectories, including a J-shaped curve for HbA1c and a U-shaped curve for LDL cholesterol. Relative to the linear model (C-index = 0.6219; Nagelkerke R² = 0.0557), both the full spline (C-index = 0.6542; R² = 0.0849; p < 0.0001) and optimised hybrid models retaining linear HbA1c (C-index = 0.6514; R² = 0.0826; p < 0.0001) substantially improved fit, discrimination and calibration. External discrimination remained superior in Holdout Set 1 (AUC = 0.6229–0.6230 vs 0.5793 linear) but attenuated in Holdout Set 2 (AUC = 0.5295–0.5354 vs 0.5231 linear).

**Conclusions:** Cardiometabolic biomarkers exhibit distinct non-linear relationships with depression in British South Asian adults, particularly within 12 months of diagnosis. Incorporating biomarker-specific functional forms significantly improves model performance over conventional linear approaches.

## Introduction

Major depressive disorder (MDD) is a heterogeneous psychiatric condition characterised by considerable variation in clinical presentation, illness course, comorbidity and treatment response [1]. Increasing evidence suggests that depression is closely associated with systemic metabolic disturbances, including altered glucose regulation, lipid metabolism, adiposity and insulin sensitivity. The relationship between depression and metabolic health is potentially bidirectional, involving shared biological mechanisms such as chronic low-grade inflammation, hypothalamic–pituitary–adrenal (HPA) axis dysregulation, altered insulin signalling and autonomic dysfunction [2]. Behavioural factors, including physical inactivity, dietary patterns and sleep disturbances, may further contribute to the co-occurrence of depressive and cardiometabolic conditions [3]. Consequently, elucidating the metabolic dimension of MDD has become a critical priority for improving risk stratification and patient outcomes.

Despite growing recognition of this metabolic dimension, associations between individual biomarkers and depression remain inconsistent. Higher body mass index (BMI), altered high-density lipoprotein (HDL) and low-density lipoprotein (LDL) cholesterol levels, elevated triglycerides, and impaired glycaemic regulation have all been implicated in depression. However, findings vary across populations and study designs [4–7]. Importantly, conventional epidemiological analyses frequently assume linear relationships between continuous biomarkers and depression. However, metabolic associations may involve threshold effects, plateaus or U-shaped relationships [8–10]. Therefore, failing to account for such non-linear dynamics represents a key methodological limitation that may obscure true dose–response patterns across studies.

A further limitation of existing research is the under-representation of South Asian populations. British Pakistani and Bangladeshi communities experience a substantial burden of cardiometabolic disease, particularly type 2 diabetes and cardiovascular conditions [11–13]. The Genes C Health (GCH) study provides an important opportunity to investigate metabolic characteristics in these populations through its large-scale, community-based recruitment and linked electronic health record (EHR) data [14, 15]. Leveraging these linked clinical records therefore provides a vital opportunity to characterise depression-related metabolic patterns directly within an under-represented, high-risk population.

In this study, we investigated metabolic characteristics associated with EHR-derived depression status in 6,340 British Bangladeshi and British Pakistani participants from the GCH cohort. We examined five metabolic and anthropometric biomarkers: BMI (Body Mass Index), HDL (High-Density Lipoprotein) cholesterol, LDL (Low-Density Lipoprotein) cholesterol, triglycerides, and glycated haemoglobin (HbA1c). Using descriptive analyses, PCA, restricted cubic splines and comparative linear, spline and optimised hybrid models, we aimed to characterise metabolic variation and evaluate whether incorporating non-linear relationships improved the modelling of depression status. Together, these analyses provide insight into population-specific metabolic profiles associated with depression in an under-represented British South Asian cohort.

## Materials and Methods

### Study Design G Population

Data were derived from the Genes C Health (GCH) longitudinal cohort of British Bangladeshi and British Pakistani adults (aged ≥16) linked with NHS primary and secondary care electronic health records (EHRs). Genetically assigned ancestry, sex, and recruitment dates were controlled for at baseline [15, 16].

### Depression Phenotype G Partitioning

Depression cases were defined using ICD-10 and SNOMED CT codes (N = 13,066), with the index date set to the first recorded diagnosis. Five cardiometabolic traits (BMI, HbA1c, HDL, LDL, triglycerides) were extracted using median values for multi-measurement windows. The primary training cohort (N = 6,340) comprised 1,585 peri-diagnostic cases matched 1:3 without replacement to 4,755 controls on age (defined as age at diagnostic event for cases and age at recruitment for controls) using nearest-neighbour Mahalanobis distance matching with a caliper of 2 units (MatchIt R package; method = “nearest”, distance = “mahalanobis”, replace = FALSE) [17]. Two independent temporal holdout sets (Set 1: N = 19,014; Set 2: N = 20,548) were structured for out-of-time generalisability testing.

### Statistical G PCA Analysis

Right-skewed continuous traits are reported as medians and IQRs, and unadjusted group comparisons are performed using two-sided Mann–Whitney U tests. Principal component analysis (PCA) was computed using the prcomp function in R to evaluate multivariate biomarker correlation structures in standardised development data.

### Regression Specifications

Multivariable logistic regression models were fitted using the rms R package [18, 19] to evaluate log-odds of EHR-derived depression status Y ∈ {0, 1} adjusted for sex and ethnicity:

1. Linear Model (7 d.f.): Models all five biomarkers linearly:

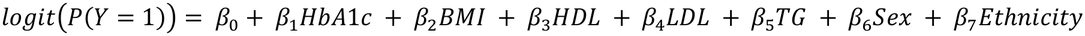
2. Full Restricted Cubic Spline Model (17 d.f.): Applies 4-knot restricted cubic splines at default quantiles (5th, 35th, 65th, 95th percentiles) to all biomarkers [20]:

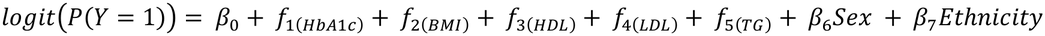 Visualisations of data distribution-anchored, adjusted odds ratios at clinical reference values (22.5 kg/m² BMI, 1.3 mmol/L HDL, 2.6 mmol/L LDL, 1.1 mmol/L triglycerides, 36 mmol/mol HbA1c).
3. Optimised Hybrid Model (15 d.f.): Retains HbA1c as a linear predictor while applying 4-knot restricted cubic splines to the remaining four cardiometabolic traits:

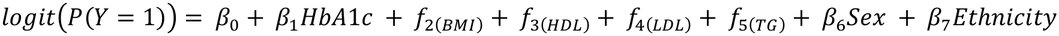

### Validation G Visualisation

Goodness-of-fit was assessed using likelihood-ratio χ² tests, Nagelkerke’s R^²^, and the C-index (AUC). Internal calibration and optimism were assessed using bootstrap resampling (rms::calibrate, B = 1,000), alongside uncalibrated external application to Holdout Sets 1 and 2. All graphics were generated using ggplot2 [21].

## Results

### Study population and metabolic characteristics

The analytical population comprised 6,340 participants from the Genes C Health cohort, including 1,585 individuals classified as depression cases and 4,755 controls based on EHR-derived depression status. Overall, the study population comprised 3,998 Bangladeshi participants (935 cases; 3,063 controls) and 2,342 Pakistani participants (650 cases; 1,692 controls) (Table 1).

**Table 1.** Unadjusted baseline metabolic and anthropometric characteristics of Genes G Health participants stratified by depression status and ethnic group (N = 6,340).

| **Biomarker / Profile** | **Cases: Bangladeshi** | **Cases: Pakistani** | **Controls: Bangladeshi** | **Controls: Pakistani** | **Overall Population** | **P-value (Bangladeshi)** | **P-value (Pakistani)** | **P-value (Overall)** |
|---|---|---|---|---|---|---|---|---|
| **Sample Size (n)** | 935 | 650 | 3063 | 1692 | 6340 |  |  |  |
| **HbA1c (mmol/mol)** | 43.00 (37.00, 53.00) | 44.00 (38.00, 58.88) | 41.50 (37.00, 50.00) | 40.50 (37.00, 48.00) | 41.50 (37.00, 51.00) | 1.879E-02 | 2.882E-12 | 4.049E-10 |
| **BMI (kg/m<sup>2</sup>)** | 26.72 (24.20, 29.78) | 29.20 (25.88, 33.04) | 26.52 (24.30, 29.20) | 28.06 (25.40, 31.12) | 27.18 (24.60, 30.30) | 1.310E-01 | 1.346E-06 | 1.708E-06 |
| **HDL Cholesterol (mmol/L)** | 1.11 (1.00, 1.35) | 1.20 (1.00, 1.40) | 1.10 (0.98, 1.30) | 1.19 (1.00, 1.40) | 1.15 (1.00, 1.35) | 2.476E-01 | 5.478E-02 | 1.303E-02 |
| **LDL Cholesterol (mmol/L)** | 2.40 (1.75, 3.10) | 2.60 (1.98, 3.15) | 2.65 (2.00, 3.28) | 2.75 (2.20, 3.20) | 2.65 (2.00, 3.20) | 3.044E-07 | 6.480E-05 | 4.057E-10 |
| **Triglycerides (mmol/L)** | 1.59 (1.15, 2.18) | 1.50 (1.07, 2.05) | 1.53 (1.15, 2.05) | 1.41 (1.05, 1.90) | 1.50 (1.11, 2.03) | 5.608E-02 | 1.157E-02 | 5.960E-03 |

*Note:* Values are presented as median (interquartile range). All comparisons are unadjusted. *P*-values were derived from two-sided Mann–Whitney *U* tests comparing depression cases and controls within the Bangladeshi subgroup, within the Pakistani subgroup, and across the overall study population. Abbreviations: BMI, body mass index; HbA1c, glycated haemoglobin; HDL, high-density lipoprotein; LDL, low-density lipoprotein.

Metabolic biomarker distributions demonstrated notable differences across depression status and ethnic groups (Table 1). Across the overall cohort, all five biomarkers differed significantly between cases and controls (p < 0.05). Within both ethnic groups, depression cases exhibited significantly higher HbA1c levels (p = 0.019 in Bangladeshi; p < 0.001 in Pakistani) and lower LDL cholesterol levels (p < 0.001 in both groups) compared with controls. In the Pakistani subgroup, depression cases also showed significantly higher BMI (p < 0.001) and triglyceride levels (p = 0.012) than controls. In contrast, differences in BMI (p = 0.131) and triglycerides (p = 0.056) did not reach statistical significance among Bangladeshi participants. HDL cholesterol did not differ significantly between cases and controls within either ethnic subgroup (p = 0.248 in Bangladeshi; p = 0.055 in Pakistani).

Across ethnicity, depression status and sex-stratified case subgroups, all biomarkers exhibited substantial inter-individual variability and marked right-skewness, with upper tails extending toward elevated values (Supplementary Figures 1–3). BMI distributions were consistently shifted toward higher values in Pakistani compared with Bangladeshi participants across both depression status and sex strata. In contrast, triglyceride concentrations were tightly concentrated centrally with pronounced right-skewed tails (Supplementary Figure 1). Similarly, HDL cholesterol showed relatively narrow distributions, while LDL cholesterol displayed a broader spread (Supplementary Figure 2), and HbA1c exhibited marked upper-tail dispersion, particularly pronounced among Pakistani depression cases (Supplementary Figure 3).

Together, these descriptive patterns highlight substantial metabolic heterogeneity across ethnic, sex and depression-status groups. Because these comparisons were unadjusted, they reflect crude observed differences rather than independent associations; moreover, the pronounced skewness and wide biomarker ranges support the evaluation of non-linear relationships alongside conventional linear models.

### Multivariate metabolic structure and principal component analysis

Principal component analysis (PCA) was performed to examine the underlying correlation structure among BMI, HDL cholesterol, LDL cholesterol, triglycerides and HbA1c. The first three principal components accounted for 72.7% of the total metabolic variance, with PC1 explaining 31.2%, PC2 explaining 22.4% and PC3 explaining 19.1% (Supplementary Table 1).

Examination of component loadings indicated distinct metabolic dimensions across the first three components (Supplementary Table 2). PC1 primarily reflected a cardiometabolic axis characterised by positive loadings for triglycerides and HbA1c (and a weak loading for BMI), opposed by negative loadings for HDL and LDL cholesterol. PC2 was driven predominantly by strong negative loadings for LDL cholesterol and BMI alongside moderate negative contributions from triglycerides, whereas PC3 was dominated by a strong negative loading for BMI with modest opposing contributions from lipid and glycaemic markers (Supplementary Table 2).

PCA biplots demonstrated extensive overlap between Bangladeshi and Pakistani participants across all pairwise combinations of the first three components (Figure 1). Although individual variation was evident across the projected metabolic space, neither ethnic group formed distinct clusters. Collectively, these findings indicate that the measured biomarkers share a largely common multivariate structure across both populations, though unsupervised PCA captures overall metabolic variance rather than depression-specific subgroups.

**Figure 1.**
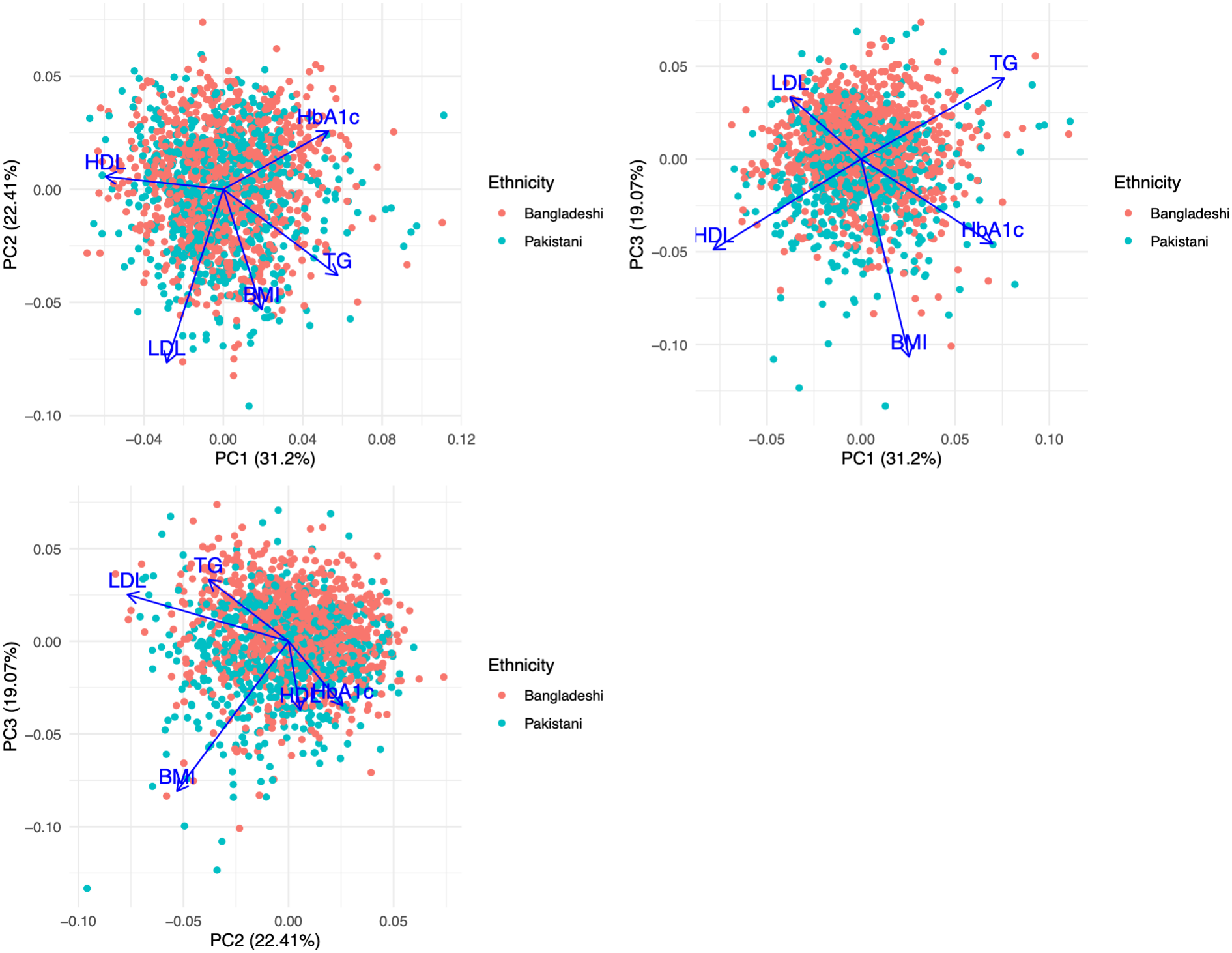
Principal component analysis (PCA) biplots of metabolic biomarkers stratified by ethnicity. Pairwise projections of the first three principal components - PC1 versus PC2 (top left), PC1 versus PC3 (top right), and PC2 versus PC3 (bottom left) - across Bangladeshi (coral) and Pakistani (teal) participants (N = 6,340). Points represent individual participant scores, and blue arrows indicate the direction and relative contribution (loadings) of body mass index (BMI), glycated haemoglobin (HbA1c), high-density lipoprotein (HDL) cholesterol, low-density lipoprotein (LDL) cholesterol, and triglycerides (TG). Percentages in axis labels denote the proportion of total variance explained by each component.

### Non-linear associations between metabolic biomarkers and depression

Multivariable restricted cubic spline analyses (with four knots per biomarker, adjusted for sex, ethnicity and all remaining biomarkers) demonstrated heterogeneous, non-linear associations between metabolic biomarkers and EHR-derived depression status (Figure 2). Across all five biomarkers, adjusted odds ratios varied across the concentration range, indicating that a single constant linear slope did not adequately capture their relationships with depression.

**Figure 2.**
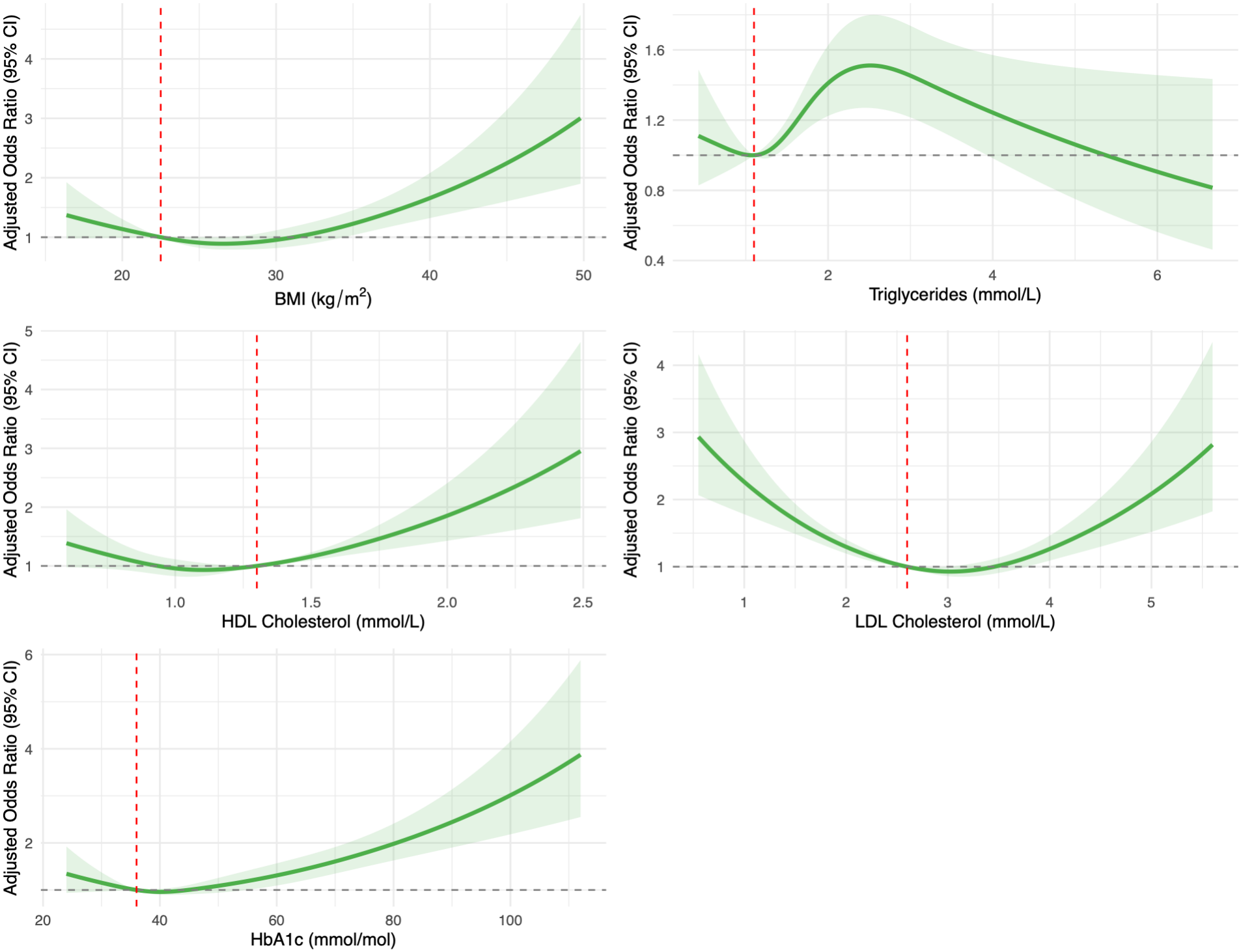
Multivariable-adjusted restricted cubic spline curves illustrating associations between continuous metabolic biomarkers and EHR-derived depression status. Solid green lines represent adjusted odds ratios and shaded green regions indicate 95% confidence intervals from a multivariable logistic regression model incorporating four-knot restricted cubic splines for body mass index (BMI, top left), triglycerides (top right), high-density lipoprotein (HDL) cholesterol (middle left), low-density lipoprotein (LDL) cholesterol (middle right), and glycated haemoglobin (HbA1c, bottom left), mutually adjusted for all other biomarkers, sex, and genetically assigned ethnicity (N = 6,340). Vertical red dashed lines indicate the clinically normal reference value set for each biomarker: 22.5 kg/m² for BMI, 1.1 mmol/L for triglycerides, 1.3 mmol/L for HDL cholesterol, 2.6 mmol/L for LDL cholesterol, and 36 mmol/mol for HbA1c (adjusted odds ratio = 1.0, denoted by the horizontal grey dashed line).

BMI showed modest variation across lower and intermediate values, followed by a steady increase in adjusted odds ratios at higher BMI levels, with widening confidence intervals at the upper tail reflecting sparser observations (Figure 2). Triglycerides exhibited a non-monotonic pattern, rising sharply above the reference level to peak at around 2.5 mmol/L before gradually declining at higher concentrations, with wide confidence intervals that warrant cautious interpretation.

HDL cholesterol showed a slight J-shaped pattern, with progressively increasing adjusted odds ratios at higher concentrations. In contrast, LDL cholesterol showed a pronounced U-shaped association, with the lowest adjusted odds ratios in the intermediate range (∼2.5–3.5 mmol/L) and elevated estimates at both extremes (Figure 2). HbA1c exhibited a clear J-shaped trajectory, remaining relatively flat below 45 mmol/mol before rising steadily across higher concentrations.

Collectively, these findings indicate substantial non-linearity across metabolic biomarkers in relation to depression status after mutual adjustment for co-occurring metabolic traits, sex and ethnicity. However, because these models did not account for additional potential confounders—such as age, medication use or socioeconomic deprivation—the curves should be interpreted as multivariable-adjusted associations rather than independent causal effects.

### Comparison of linear and restricted cubic spline models

To evaluate whether modelling non-linear biomarker trajectories improved predictive performance, a conventional multivariable linear logistic regression model was compared with a full four-knot restricted cubic spline model, with both frameworks adjusted for sex and genetically assigned ethnicity. Incorporating non-linear spline terms substantially improved both goodness-of-fit and discriminative accuracy relative to the linear specification. Specifically, compared with the multivariable linear model (7 degrees of freedom; C-index = 0.6219; Nagelkerke R² = 0.0557), the full restricted cubic spline model (17 degrees of freedom) significantly improved overall model fit (likelihood-ratio χ² = 131.29, p < 0.0001) and increased both discrimination (C-index = 0.6542; ΔC-index = 0.0323) and explained variance (Nagelkerke R² = 0.0849; ΔR² = 0.0292). These findings demonstrate that accounting for non-linear biomarker relationships significantly enhances the characterisation and discrimination of EHR-derived depression status compared with conventional linear modelling.

### Relative predictor importance and optimised hybrid model

Variable importance analysis -quantified by partial Wald χ² minus degrees of freedom - demonstrated distinct shifts in predictor contributions between the linear and full restricted cubic spline specifications (Figure 3, top panel). Across both models, gender was the strongest individual predictor, whereas ethnicity contributed minimally. Among the metabolic biomarkers, HbA1c was the dominant contributor in the linear model, followed by LDL cholesterol, HDL cholesterol, triglycerides and BMI. Incorporating four-knot restricted cubic splines substantially increased the partial contributions of LDL cholesterol (which became the leading metabolic predictor), BMI, HDL cholesterol and triglycerides. In contrast, the partial Wald χ² - d.f. of HbA1c decreased under the spline specification compared with its linear parameterisation.

**Figure 3.**
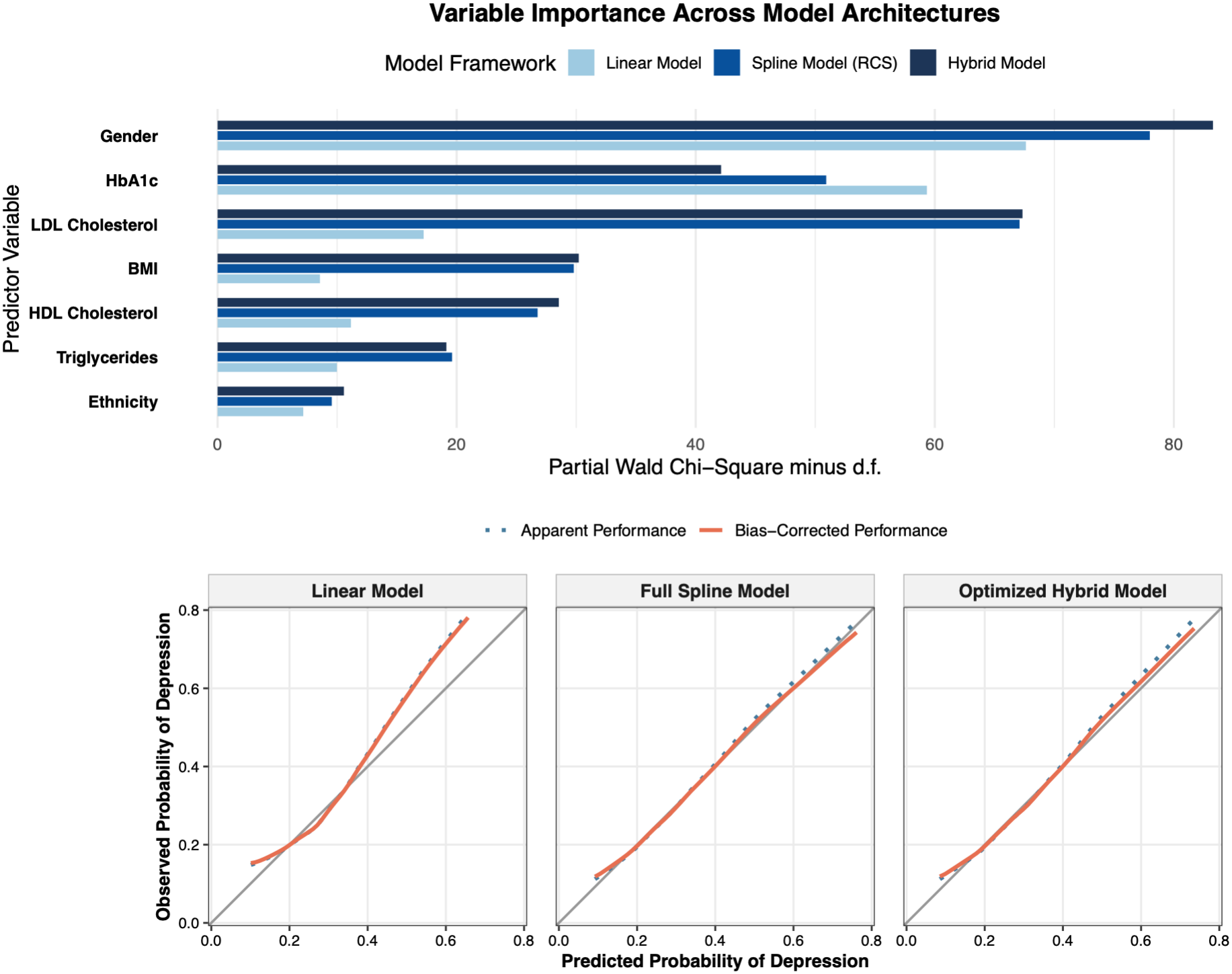
Relative predictor importance and bootstrap calibration across multivariable linear, full spline, and optimised hybrid models. Top: Variable importance quantified by partial Wald chi-square minus degrees of freedom (χ² - d.f.) across predictors in the multivariable linear model (light blue), full four-knot restricted cubic spline model (medium blue), and optimised hybrid model retaining glycated haemoglobin (HbA1c) as a linear term (dark blue). Bottom: Apparent (dotted blue) and bootstrap bias-corrected (solid orange) calibration curves comparing predicted versus observed probabilities of EHR-derived depression status against the ideal calibration diagonal (grey line) for the linear (left), full spline (centre), and optimised hybrid (right) models in the Genes C Health cohort (N = 6,340). Abbreviations: BMI, body mass index; d.f., degrees of freedom; EHR, electronic health record; HbA1c, glycated haemoglobin; HDL, high-density lipoprotein; LDL, low-density lipoprotein.

Guided by this variable importance profile, we evaluated an optimised hybrid framework in which HbA1c was retained as a linear term. At the same time, BMI, HDL cholesterol, LDL cholesterol and triglycerides were modelled using four-knot restricted cubic splines (Figure 3). Reducing model complexity from 17 to 15 degrees of freedom, the optimised hybrid model significantly outperformed the conventional multivariable linear model (likelihood-ratio χ² = 120.75, p < 0.0001), achieving a Nagelkerke R^2^ of 0.0826 (ΔR² = 0.0269) and a C-index of 0.6514 (ΔC-index = 0.0295) and notably, retaining HbA1c as a linear predictor preserved nearly all of the goodness-of-fit and discriminative gains of the full four-knot spline model (R^2^ = 0.0849; C-index = 0.6542) within a more parsimonious model architecture. These importance metrics reflect relative statistical contributions within each specified model rather than independent causal effects.

### Calibration and comparative interquartile associations

Bootstrap calibration analysis demonstrated close agreement between apparent and bias-corrected calibration curves across all three frameworks, indicating minimal overfitting (Figure 3, bottom panel). However, the linear model showed noticeable departure from the ideal diagonal reference line -particularly at predicted probabilities below 0.20 and above 0.40. In contrast, both the full spline and optimised hybrid models tracked the ideal calibration line closely across the prediction range, confirming that reducing complexity in the hybrid framework did not compromise calibration accuracy.

Comparative interquartile odds ratios (comparing the 75th versus 25th percentiles of each continuous biomarker) further highlighted differences in estimated associations across modelling frameworks (Supplementary Figure 4). LDL cholesterol demonstrated a consistent inverse interquartile association with depression across all three models, with substantially stronger inverse estimates in the spline and hybrid frameworks than in the linear model. Conversely, triglycerides exhibited a markedly stronger positive interquartile association under the spline and hybrid specifications compared with the linear model. For HbA1c, positive interquartile associations were clearly preserved with narrow confidence intervals in both the linear and hybrid models. In contrast, the four-knot spline estimate was attenuated, with a confidence interval spanning unity, further supporting the retention of HbA1c as a linear term in the hybrid framework. By contrast, HDL cholesterol and BMI showed modest positive interquartile associations in the linear model, which attenuated in the spline and hybrid frameworks, with confidence intervals crossing unity, consistent with their non-linear risk profiles at extreme values (Supplementary Figure 4). Gender and ethnicity remained consistently associated with depression status across all three modelling architectures.

### Temporal validation in independent holdout datasets

To evaluate temporal generalisability without data leakage, all three trained models were applied without recalibration to two independent holdout datasets comprising participants completely non-overlapping with the development cohort. Both holdout sets were defined by the timing of metabolic measurements relative to the first recorded depression event and evaluated against a shared independent control group (n = 18,733: 12,407 Bangladeshi and 6,326 Pakistani; Supplementary Tables 3–4). Holdout Set 1 (N = 19,014) comprised 281 non-overlapping cases (182 Bangladeshi, 99 Pakistani; prevalence 1.48%) with metabolic measurements recorded 6 to 12 months before their first depression event, whereas Holdout Set 2 (N = 20,548) comprised 1,815 non-overlapping cases (1,260 Bangladeshi, 555 Pakistani; prevalence 8.83%) with measurements recorded more than 12 months before their first depression event.

Baseline biomarker profiles varied markedly with proximity to the onset of depression (Supplementary Tables 3–4). In Holdout Set 1 (6–12 months pre-onset), metabolic patterns closely mirrored those of the primary development cohort, with cases showing modestly elevated HbA1c and BMI alongside lower LDL cholesterol levels relative to shared controls (Supplementary Table 3). By contrast, in Holdout Set 2 (>12 months pre-onset), this metabolic contrast was absent or reversed, with cases exhibiting slightly lower median HbA1c and triglyceride levels than controls (Supplementary Table 4).

Consistent with these temporal biomarker profiles, discriminative performance was strongest near the onset of depression. In Holdout Set 1 (6–12 months pre-onset), both the full restricted cubic spline model (external AUC = 0.6229) and the optimised hybrid framework (external AUC = 0.6230) substantially outperformed the multivariable linear baseline (external AUC = 0.5793), yielding incremental AUC gains of 0.0436 and 0.0437, respectively (Figure 4). In Holdout Set 2 (>12 months pre-onset), overall discrimination attenuated across all architectures. However, both the full spline (external AUC = 0.5354) and optimised hybrid (external AUC = 0.5295) models continued to exceed the linear baseline (external AUC = 0.5231; Figure 5).

**Figure 4.**
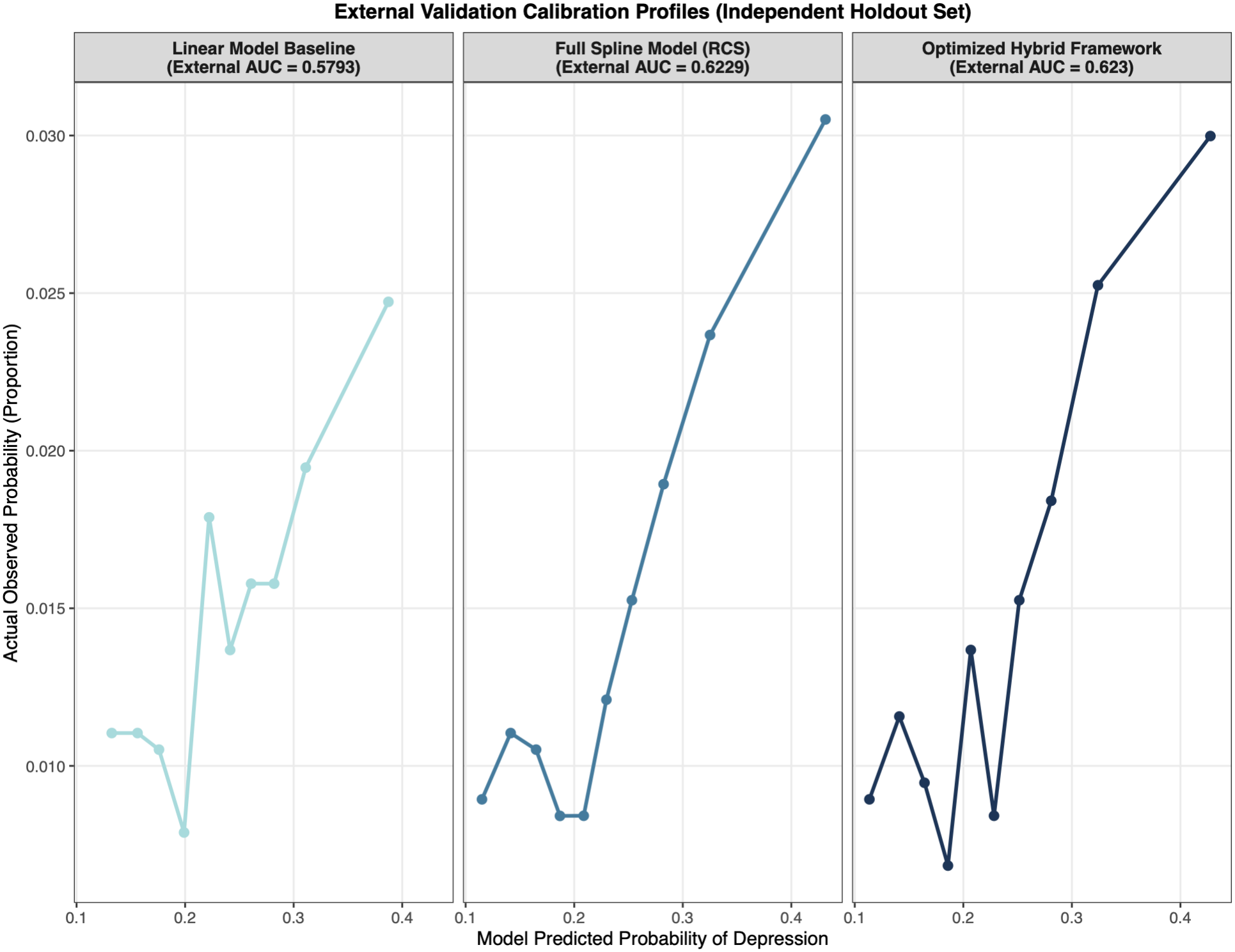
External validation calibration profiles in Holdout Set 1 (non-overlapping cases measured 6 to 12 months before first depression event). Decile-based calibration curves comparing model-predicted probabilities against actual observed proportions of EHR-derived depression across the multivariable linear baseline (left, light blue), full four-knot restricted cubic spline model (centre, medium blue), and optimised hybrid framework retaining linear HbA1c (right, dark blue). Cases were independent of the development cohort, defined by metabolic measurements recorded 6 to 12 months before age at first recorded depression event (n = 281: 182 Bangladeshi, 99 Pakistani), and evaluated alongside shared independent controls (n = 18,733: 12,407 Bangladeshi, 6,326 Pakistani; total N = 19,014). External area under the receiver operating characteristic curve (AUC) values are indicated in each panel header.

**Figure 5.**
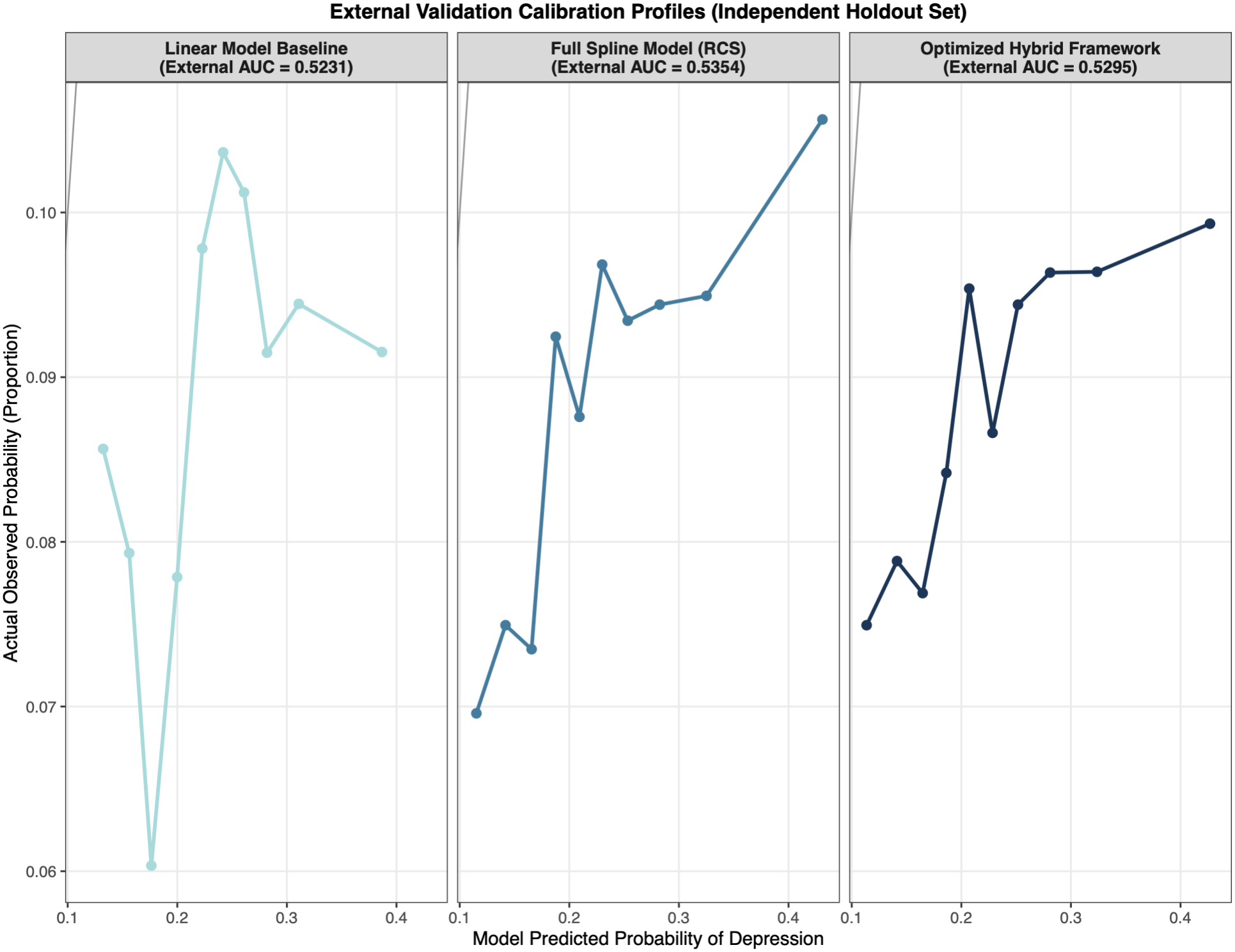
External validation calibration profiles in Holdout Set 2 (non-overlapping cases measured more than 12 months before first depression event). Decile-based calibration curves comparing model-predicted probabilities against actual observed proportions of EHR-derived depression across the multivariable linear baseline (left, light blue), full four-knot restricted cubic spline model (centre, medium blue), and optimised hybrid framework retaining linear HbA1c (right, dark blue). Cases were independent of the development cohort, defined by metabolic measurements recorded more than 12 months before age at first recorded depression event (n = 1,815: 1,260 Bangladeshi, 555 Pakistani), and evaluated alongside shared independent controls (n = 18,733: 12,407 Bangladeshi, 6,326 Pakistani; total N = 20,548). External area under the receiver operating characteristic curve (AUC) values are indicated in each panel header.

Across both holdout evaluations, model-predicted probabilities (∼0.10–0.45) exceeded actual observed proportions (∼0.007–0.031 in Holdout Set 1 and ∼0.060–0.106 in Holdout Set 2), reflecting the lower case prevalence in the holdout sets compared with the 25.0% prevalence in the development cohort (Figures 4–5). Nevertheless, calibration profiles demonstrated superior monotonic risk stratification across higher predicted-risk deciles for the full spline and optimised hybrid frameworks, compared with the erratic decile trajectory of the linear baseline.

## Discussion

In this study, we investigated metabolic characteristics associated with EHR-derived depression status among 6,340 British Bangladeshi and British Pakistani participants in the Genes C Health cohort, followed by temporal validation in two independent holdout datasets. Four principal findings emerged. First, depression cases differed significantly from controls across several metabolic traits - most consistently exhibiting higher median HbA1c and lower LDL cholesterol across both ethnic groups, alongside higher BMI and triglycerides among Pakistani participants. Second, multivariable restricted cubic spline analyses revealed heterogeneous, non-linear relationships across biomarkers, including a J-shaped trajectory for HbA1c, a U-shaped association for LDL cholesterol, a non-monotonic trajectory for triglycerides and upward tail trends for BMI and HDL cholesterol. Third, incorporating non-linear spline terms substantially improved model fit and discrimination relative to a conventional linear specification, while an optimised hybrid framework retaining HbA1c as a linear term preserved virtually identical performance with reduced model complexity. Fourth, in independent temporal holdout evaluations, both non-linear models maintained superior discrimination over the linear baseline, particularly when metabolic measurements were recorded within 6 to 12 months prior to the first recorded depression event.

### Metabolic heterogeneity and lipid–glycaemic patterns

The consistent elevation of HbA1c among depression cases in both Bangladeshi and Pakistani participants aligns with extensive epidemiological evidence linking depression with insulin resistance and impaired glycaemic regulation [7, 22]. Shared biological mechanisms - including hypothalamic–pituitary–adrenal axis dysregulation, chronic low-grade inflammation and altered insulin signalling - alongside behavioural factors and psychotropic medication exposure likely contribute to this co-occurrence [23–25]. Although BMI differences were more pronounced in Pakistani than Bangladeshi participants, substantial inter-individual dispersion across all biomarkers highlights the importance of evaluating multi-domain metabolic profiles rather than single indicators alone.

Notably, our observation of lower median LDL cholesterol and an inverse interquartile association between LDL cholesterol and depression corroborates recent large-scale prospective findings from the UK Biobank, which demonstrated robust inverse associations between LDL cholesterol, apolipoprotein B and total cholesterol and incident depression across 445,105 adults [26]. The report highlighted that systemic cholesterol availability plays a vital neurobiological role in maintaining neuronal membrane fluidity, myelin integrity, and synaptic receptor function, such that lower circulating LDL cholesterol may reflect compromised lipid trafficking, increased neurobiological vulnerability, or, in clinical EHR cohorts, higher rates of lipid-lowering therapy among individuals with cardiometabolic comorbidities. At the same time, our spline analysis extended this observation by revealing a U-shaped trajectory for LDL cholesterol, indicating that both very low and very high concentrations may be associated with elevated odds of depression, consistent with prior non-linear reports [27–29].

### Multivariate architecture and non-linear hybrid modelling

Principal component analysis revealed an interconnected multivariate metabolic architecture, with the first three components explaining 72.7% of the total variance, anchored by a primary cardiometabolic axis contrasting triglycerides and HbA1c with HDL and LDL cholesterol. Extensive overlap between Bangladeshi and Pakistani participants across PCA biplots indicates that the underlying correlation structure among these biomarkers is largely shared across both South Asian subgroups, even though unsupervised components do not separate depression cases from controls.

In supervised multivariable modelling, replacing rigid linear assumptions with four-knot restricted cubic splines significantly improved both goodness-of-fit and discrimination (C-index increasing from 0.6219 to 0.6542) and corrected the calibration deviations observed in the linear model. Variable importance profiling further revealed that while spline parameterisation markedly increased the explanatory contributions of LDL cholesterol, BMI, HDL cholesterol and triglycerides, HbA1c retained stronger penalty-adjusted importance as a linear predictor. Guided by this insight, our optimised hybrid framework - combining linear HbA1c with spline-transformed lipid and adiposity markers - achieved virtually identical discrimination (C-index = 0.6514) and calibration accuracy with fewer degrees of freedom. These findings align with recent UK Biobank evidence demonstrating that biomarker-specific functional forms are essential for modelling depression risk, as uniform linear specifications can obscure threshold effects and U-shaped associations at the extremes of the distribution [26].

### Temporal generalisability across independent holdout cohorts

External validation in two independent, non-overlapping holdout datasets provided important insight into the temporal dynamics of metabolic dysregulation preceding depression diagnosis. When metabolic biomarkers were measured 6 to 12 months before the first recorded depression event (Holdout Set 1), biomarker contrasts mirrored the development cohort and both the full spline (external AUC = 0.6229) and optimised hybrid (external AUC = 0.6230) models outperformed the linear baseline (external AUC = 0.5793) by more than 0.043 AUC units. By contrast, when measurements were recorded more than 12 months before the onset of depression (Holdout Set 2), metabolic differences between cases and controls attenuated, and overall discrimination declined across all architectures, though non-linear models remained superior. This temporal gradient suggests that metabolic perturbations - particularly dysglycaemia and dyslipidaemia - may intensify during the prodromal window preceding clinical recognition of depression, whereas measurements taken further in advance capture weaker preclinical divergence [30–32]. Although absolute predicted probabilities exceeded observed proportions due to lower case prevalence in the holdout datasets compared with the 25.0% development sample, both non-linear frameworks preserved superior monotonic risk stratification across higher risk deciles.

### Strengths and limitations

Key strengths of this study include its focus on an under-represented, high-cardiometabolic-risk British Bangladeshi and Pakistani population; the integration of routine EHR biomarkers with genetically assigned ethnicity; the rigorous comparison of linear, full-spline, and parsimonious hybrid parameterisations; and validation across temporally stratified, non-overlapping holdout datasets. Nevertheless, several limitations warrant consideration. First, although multivariable models adjusted for sex, ethnicity and co-occurring metabolic biomarkers, they did not account for age, socioeconomic deprivation, smoking status, psychotropic or lipid-lowering medications (such as statins and antidepressants), or pre-existing cardiometabolic diagnoses; consequently, residual confounding cannot be excluded, and findings should be interpreted as multivariable associations rather than causal effects. Second, EHR-derived depression status reflects clinical recognition and coding in primary or secondary care, which may under-ascertain milder or untreated cases and introduce timing imprecision between biological onset and recorded diagnosis. Third, biomarkers were analysed using single time-point measurements rather than longitudinal within-person trajectories. Finally, because Genes C Health participants are recruited from specific UK regions, generalisability to other South Asian diaspora or global populations requires further evaluation.

## Conclusions

In summary, metabolic biomarkers exhibit complex, non-linear relationships with EHR-derived depression status in British Bangladeshi and British Pakistani adults. Incorporating biomarker-specific functional forms through restricted cubic splines, or a parsimonious hybrid architecture that combines linear HbA1c with non-linear lipid and adiposity terms, substantially improves model fit, calibration, and temporal discrimination compared with conventional linear approaches. Future prospective studies incorporating longitudinal biomarker trajectories, medication records and comprehensive sociodemographic covariates are warranted to clarify causal mechanisms and evaluate whether non-linear metabolic profiling can support early risk stratification in high-risk clinical populations.

## Ethics Declarations

Genes C Health was approved by the National Research Ethics Committee (London and Southeast), and the Health Research Authority (reference 13/LO/124). Written informed consent authorising the collection, analysis, and publication of health and genetic data was obtained from all study volunteers.

## Data Availability

All summary-level data supporting the findings of this study are available within the article and its supplementary information. Owing to participant privacy and governance restrictions, individual-level participant data cannot be shared publicly and are accessible only within the Genes C Health Trusted Research Environment (TRE) under controlled access. Bona fide researchers may apply for data access subject to approval by the Genes C Health Executive Committee and execution of a Data Access Agreement. Full application details are available at https://www.genesandhealth.org/researchers/apply-for-access/.

## Acknowledgment

Genes C Health is/has recently been core-funded by Wellcome (WT102627, WT210561), the Medical Research Council (UK) (M009017, MR/X009777/1, MR/X009920/1), Higher Education Funding Council for England Catalyst, Barts Charity (845/1796), Health Data Research UK (for London substantive site), and research delivery support from the NHS National Institute for Health Research Clinical Research Network (North Thames). We acknowledge the support of the National Institute for Health and Care Research Barts Biomedical Research Centre (NIHR203330); a delivery partnership of Barts Health NHS Trust, Queen Mary University of London, St George’s University Hospitals NHS Foundation Trust and St George’s University of London

Genes C Health is/has recently been funded by Alnylam Pharmaceuticals, Genomics PLC; and a Life Sciences Industry Consortium of AstraZeneca PLC, Bristol-Myers Squibb Company, GlaxoSmithKline Research and Development Limited, Maze Therapeutics Inc, Merck Sharp C Dohme LLC, Novo Nordisk A/S, Pfizer Inc, Takeda Development Centre Americas Inc.

We thank Social Action for Health, Centre of The Cell, members of our Community Advisory Group, and staff who have recruited and collected data from volunteers. We thank the NIHR National Biosample Centre (UK Biocentre), the Social Genetic C Developmental Psychiatry Centre (King’s College London), Wellcome Sanger Institute, and Broad Institute for sample processing, genotyping, sequencing and variant annotation. This work uses data provided by patients and collected by the NHS as part of their care and support. This research utilised Queen Mary University of London’s Apocrita HPC facility, supported by QMUL Research-IT, http://doi.org/10.5281/zenodo.438045

We thank: Barts Health NHS Trust, NHS Clinical Commissioning Groups (City and Hackney, Waltham Forest, Tower Hamlets, Newham, Redbridge, Havering, Barking and Dagenham), East London NHS Foundation Trust, Bradford Teaching Hospitals NHS Foundation Trust, Public Health England (especially David Wyllie), Discovery Data Service/Endeavour Health Charitable Trust (especially David Stables), Voror Health Technologies Ltd (especially Sophie Don), NHS England (for what was NHS Digital) - for GDPR-compliant data sharing backed by individual written informed consent.

Most of all we thank all of the volunteers participating in Genes C Health.

A favourable ethical opinion for the main Genes C Health research study was granted by NRES Committee London - South East (reference 14/LO/1240) on 16 Sept 2014. Queen Mary University of London is the Sponsor, and Data Controller.

## Supplementary Figures and Tables

**Supplementary Figure 1**. Distributions of body mass index (BMI) and triglycerides (TG) by depression status, sex, and ethnicity in the Genes C Health cohort (N = 6,340).

**Supplementary Figure 2.** Distributions of high-density lipoprotein (HDL) and low-density lipoprotein (LDL) cholesterol by depression status, sex, and ethnicity in the Genes C Health cohort (N = 6,340).

**Supplementary Figure 3.** Distribution of glycated haemoglobin (HbA1c) by depression status, sex, and ethnicity in the Genes C Health cohort (N = 6,340).

**Supplementary Figure 4.** Comparative interquartile odds ratios (95% confidence intervals) across linear, full spline, and optimised hybrid models. Forest plot displaying multivariable-adjusted odds ratios (log scale) and 95% confidence intervals for EHR-derived depression status across the multivariable linear model (light blue), full four-knot restricted cubic spline model (medium blue), and optimised hybrid model with linear HbA1c (dark blue) in the Genes C Health cohort (N = 6,340). For continuous metabolic biomarkers—low-density lipoprotein (LDL) cholesterol, glycated haemoglobin (HbA1c), high-density lipoprotein (HDL) cholesterol, triglycerides, and body mass index (BMI)—odds ratios represent interquartile comparisons (75th versus 25th percentile). Categorical predictors represent sex (Gender) and genetically assigned ethnicity (Ethnicity [PCA]). The vertical red dashed line denotes the null effect (odds ratio = 1.0).

**Supplementary Table 1:** Standard deviation, proportion of variance explained, and cumulative proportion of variance across all five principal components derived from metabolic biomarkers.

| Metric | PC1 | PC2 | PC3 | PC4 | PC5 |
|---|---|---|---|---|---|
| Standard deviation | 1.249 | 1.059 | 0.977 | 0.884 | 0.765 |
| Proportion of Variance | 0.312 | 0.224 | 0.191 | 0.156 | 0.117 |
| Cumulative Proportion | 0.312 | 0.536 | 0.727 | 0.883 | 1 |

**Supplementary Table 2:** Variable loadings for the first three principal components (PC1–PC3) across body mass index (BMI), high-density lipoprotein (HDL) cholesterol, low-density lipoprotein (LDL) cholesterol, triglycerides (TG), and glycated haemoglobin (HbA1c).

| Biomarker | PC1 | PC2 | PC3 |
|---|---|---|---|
| BMI | 0.186 | -0.51 | -0.776 |
| HDL | -0.571 | 0.053 | -0.356 |
| LDL | -0.274 | -0.737 | 0.242 |
| TG | 0.554 | -0.365 | 0.319 |
| HbA1c | 0.508 | 0.247 | -0.333 |

**Supplementary Table 3:** Baseline metabolic and anthropometric characteristics of Holdout Set 1 participants (cases measured 6 to 12 months before first depression event) stratified by depression status and ethnic group (N = 19,014).

| Biomarker / Profile | Cases: Bangladeshi | Cases: Pakistani | Controls: Bangladeshi | Controls: Pakistani | Overall Population |
|---|---|---|---|---|---|
| Sample Size (n) | 182 | 99 | 12407 | 6326 | 19014 |
| HbA1c (mmol/mol) | 41.00 (37.00, 50.75) | 39.00 (35.00, 43.09) | 40.00 (36.00, 47.00) | 39.00 (36.00, 44.00) | 39.50 (36.00, 46.00) |
| BMI (kg/m<sup>2</sup>) | 26.97 (24.15, 30.21) | 28.09 (24.90, 31.00) | 26.53 (24.28, 29.29) | 27.75 (24.90, 31.14) | 26.90 (24.43, 29.90) |
| HDL Cholesterol (mmol/L) | 1.14 (1.00, 1.30) | 1.19 (1.00, 1.32) | 1.13 (1.00, 1.30) | 1.20 (1.01, 1.40) | 1.15 (1.00, 1.35) |
| LDL Cholesterol (mmol/L) | 2.50 (1.80, 3.18) | 2.60 (2.17, 3.30) | 2.70 (2.15, 3.30) | 2.71 (2.24, 3.25) | 2.70 (2.20, 3.30) |
| Triglycerides (mmol/L) | 1.59 (1.17, 2.27) | 1.29 (0.98, 2.12) | 1.52 (1.11, 2.10) | 1.35 (0.98, 1.84) | 1.46 (1.07, 2.02) |

Note: Values are presented as median (interquartile range). Cases comprise participants with metabolic measurements recorded 6 to 12 months before age at first recorded depression event (n = 281), evaluated against shared external controls without recorded depression (n = 18,733). Abbreviations: BMI, body mass index; HbA1c, glycated haemoglobin; HDL, high-density lipoprotein; LDL, low-density lipoprotein.

**Supplementary Table 4:** Baseline metabolic and anthropometric characteristics of Holdout Set 2 participants (cases measured more than 12 months before first depression event) stratified by depression status and ethnic group (N = 20,548).

| Biomarker / Profile | Cases: Bangladeshi | Cases: Pakistani | Controls: Bangladeshi | Controls: Pakistani | Overall Population |
|---|---|---|---|---|---|
| Sample Size (n) | 1260 | 555 | 12407 | 6326 | 20548 |
| HbA1c (mmol/mol) | 39.00 (35.50, 45.00) | 37.00 (34.00, 41.00) | 40.00 (36.00, 47.00) | 39.00 (36.00, 44.00) | 39.50 (36.00, 46.00) |
| BMI (kg/m<sup>2</sup>) | 26.60 (24.00, 29.50) | 27.10 (23.92, 31.28) | 26.53 (24.28, 29.29) | 27.75 (24.90, 31.14) | 26.90 (24.40, 29.90) |
| HDL Cholesterol (mmol/L) | 1.15 (1.00, 1.37) | 1.23 (1.05, 1.40) | 1.13 (1.00, 1.30) | 1.20 (1.01, 1.40) | 1.16 (1.00, 1.35) |
| LDL Cholesterol (mmol/L) | 2.80 (2.20, 3.30) | 2.70 (2.29, 3.23) | 2.70 (2.15, 3.30) | 2.71 (2.24, 3.25) | 2.70 (2.20, 3.30) |
| Triglycerides (mmol/L) | 1.43 (1.04, 2.00) | 1.18 (0.88, 1.69) | 1.52 (1.11, 2.10) | 1.35 (0.98, 1.84) | 1.45 (1.06, 2.00) |

Note: Values are presented as median (interquartile range). Cases comprise participants with metabolic measurements recorded more than 12 months before age at first recorded depression event (n = 1,815), evaluated against shared external controls without recorded depression (n = 18,733). Abbreviations: BMI, body mass index; HbA1c, glycated haemoglobin; HDL, high-density lipoprotein; LDL, low-density lipoprotein.

