## Supplementary Figures for "The Non-Linear Metabolic Architecture of Depression Risk: A Cross-Phenotypic Hybrid Modelling Analysis in Genes and Health Cohort"

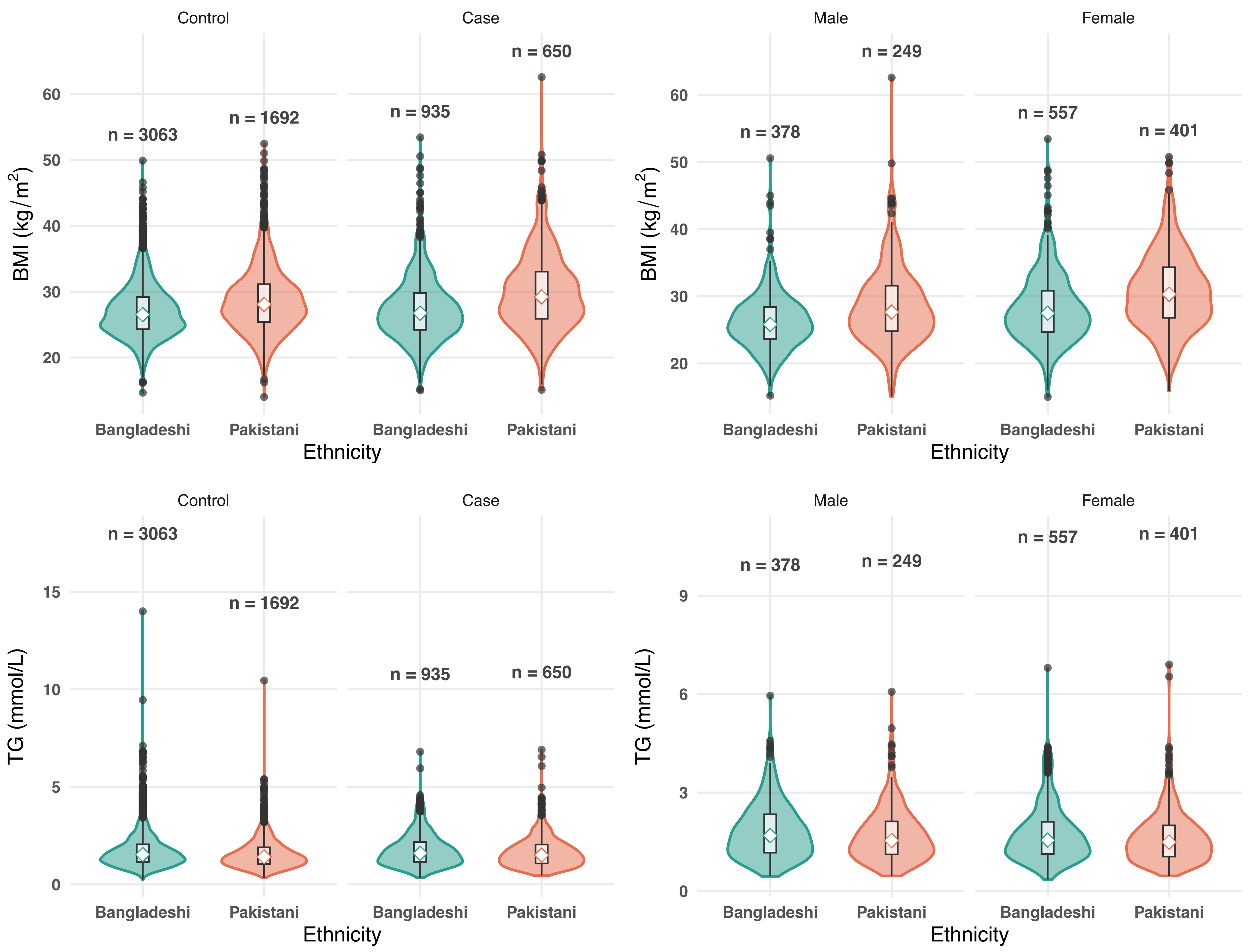

Control
Case
Male
Female
BMI (kg/m2)
TG (mmol/L)
n = 3063
n = 1692
n = 935
n = 650
n = 378
n = 249
n = 557
n = 401
Bangladeshi
Pakistani
Ethnicity

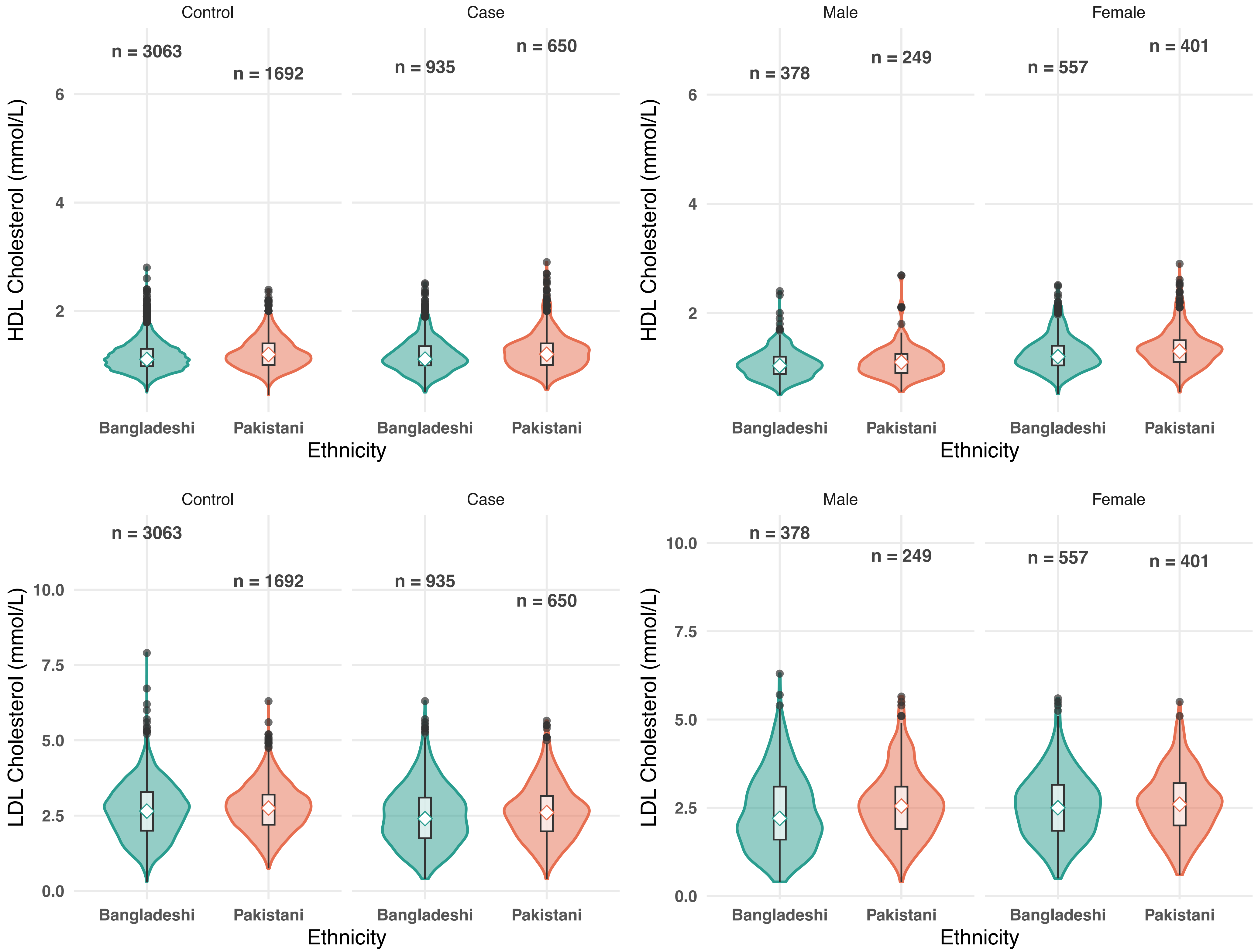
Control
Case
Male
Female
n = 3063
n = 1692
n = 935
n = 650
n = 378
n = 249
n = 557
n = 401
HDL Cholesterol (mmol/L)
6
4
2
Bangladeshi
Pakistani
Ethnicity
LDL Cholesterol (mmol/L)
10.0
7.5
5.0
2.5
0.0

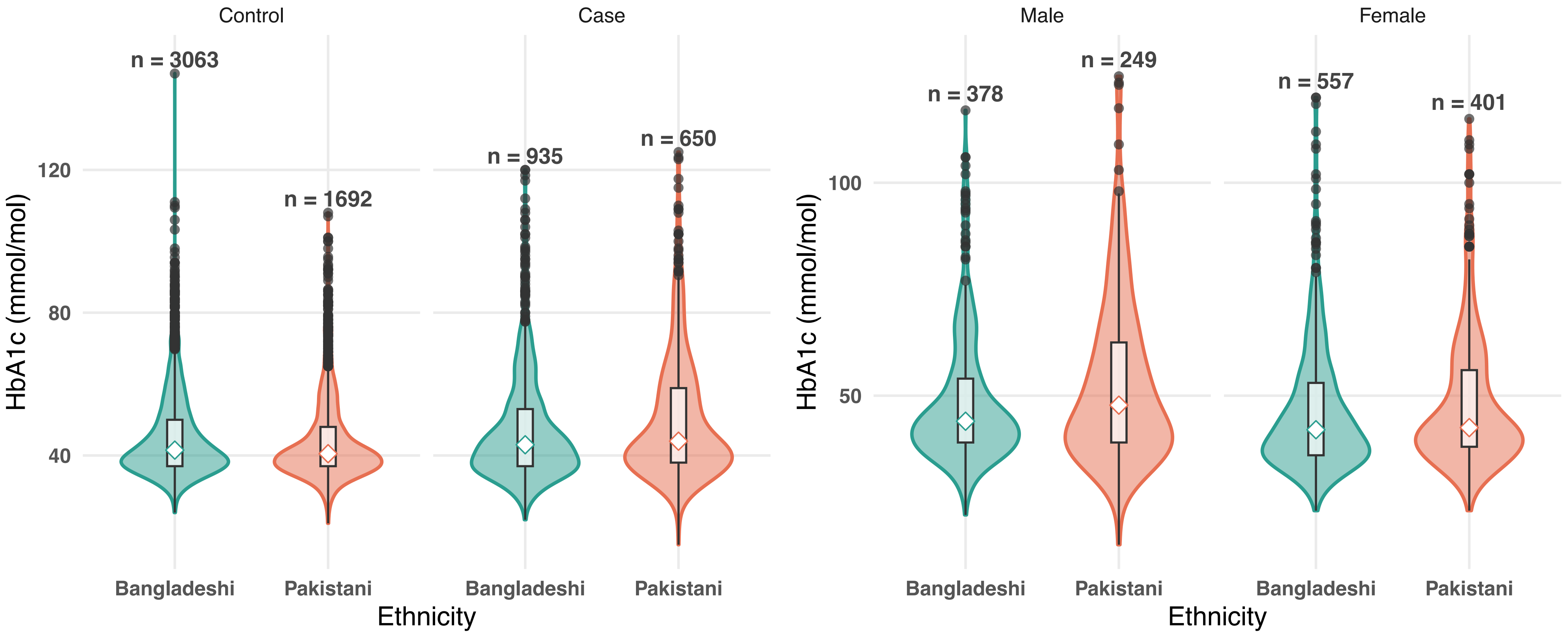

Control
Case
Male
Female
n = 3063
n = 1692
n = 935
n = 650
n = 378
n = 249
n = 557
n = 401
HbA1c (mmol/mol)
120
80
40
100
50
Bangladeshi
Pakistani
Ethnicity

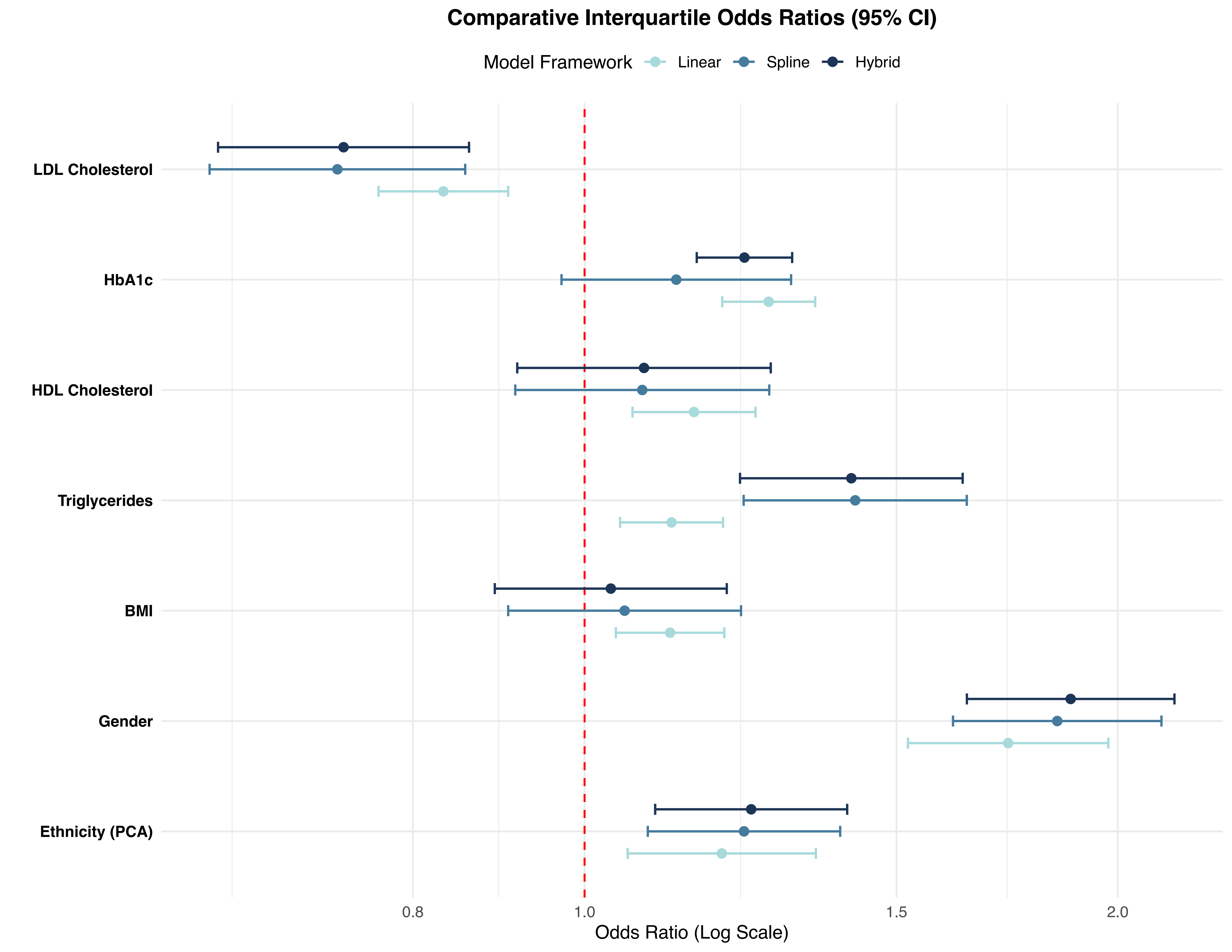

Comparative Interquartile Odds Ratios (95% CI)
Model Framework
Linear
Spline
Hybrid
LDL Cholesterol
HbA1c
HDL Cholesterol
Triglycerides
BMI
Gender
Ethnicity (PCA)
0.8
1.0
1.5
2.0
Odds Ratio (Log Scale)
